# Detection and surveillance of SARS-CoV-2 genomic variants in wastewater

**DOI:** 10.1101/2021.01.08.21249379

**Authors:** Katharina Jahn, David Dreifuss, Ivan Topolsky, Anina Kull, Pravin Ganesanandamoorthy, Xavier Fernandez-Cassi, Carola Bänziger, Alexander J. Devaux, Elyse Stachler, Lea Caduff, Federica Cariti, Alex Tuñas Corzón, Lara Fuhrmann, Chaoran Chen, Kim Philipp Jablonski, Sarah Nadeau, Mirjam Feldkamp, Christian Beisel, Catharine Aquino, Tanja Stadler, Christoph Ort, Tamar Kohn, Timothy R. Julian, Niko Beerenwinkel

**Affiliations:** Department of Biosystems Science and Engineering, ETH Zurich, CH-4058 Basel, Switzerland; SIB Swiss Institute of Bioinformatics, CH-1015 Lausanne, Switzerland; Eawag, Swiss Federal Institute of Aquatic Science and Technology, CH-8600 Dübendorf, Switzerland; Laboratory of Environmental Chemistry, School of Architecture, Civil and Environmental Engineering, École Polytechnique Fédérale de Lausanne (EPFL), CH-1015 Lausanne, Switzerland; Functional Genomics Center Zurich, ETH Zurich, CH-8057 Zurich, Switzerland; Swiss Tropical and Public Health Institute, CH-4051 Basel, Switzerland; University of Basel, CH-4055 Basel, Switzerland

## Abstract

The emergence of SARS-CoV-2 mutants with altered transmissibility, virulence, or immunogenicity emphasizes the need for early detection and epidemiological surveillance of genomic variants. Wastewater samples provide an opportunity to assess circulating viral lineages in the community. We performed genomic sequencing of 122 wastewater samples from three locations in Switzerland to analyze the B.1.1.7, B.1.351, and P.1 variants of SARS-CoV-2 on a population level. We called variant-specific signature mutations and monitored variant prevalence in the local population over time. To enable early detection of emerging variants, we developed a bioinformatics tool that uses read pairs carrying multiple signature mutations as a robust indicator of low-frequency variants. We further devised a statistical approach to estimate the transmission fitness advantage, a key epidemiological parameter indicating the speed at which a variant spreads through the population, and compared the wastewater-based findings to those derived from clinical samples. We found that the local outbreak of the B.1.1.7 variant in two Swiss cities was observable in wastewater up to 8 days before its first detection in clinical samples. We detected a high prevalence of the B.1.1.7 variant in an alpine ski resort popular among British tourists in December 2020, a time when the variant was still very rare in Switzerland. We found no evidence of local spread of the B.1.351 and P.1 variants at the monitored locations until the end of the study (mid February) which is consistent with clinical samples. Estimation of local variant prevalence performs equally well or better for wastewater samples as for a much larger number of clinical samples. We found that the transmission fitness advantage of B.1.1.7, i.e. the relative change of its reproductive number, can be estimated earlier and based on substantially fewer wastewater samples as compared to using clinical samples. Our results show that genomic sequencing of wastewater samples can detect, monitor, and evaluate genetic variants of SARS-CoV-2 on a population level. Our methodology provides a blueprint for rapid, unbiased, and cost-efficient genomic surveillance of SARS-CoV-2 variants.

## Introduction

The ongoing spread and evolution of SARS-CoV-2 has generated several variants of interest and variants of concern (1–3), which can affect, to different degrees, transmissibility (1), disease severity (4), diagnostics, and the effectiveness of treatment (5) and vaccines. Therefore, early detection and monitoring local variant spread has become an important public health task (6).

Viral RNA of SARS-CoV-2 infected persons can be detected in the sewage collected in wastewater treatment plants (WWTPs) and its concentration has been shown to correlate with case reports (7). Moreover, wastewater samples can provide a snapshot of the circulating viral lineages and their diversity in the community through RT-qPCR analysis (8,9) or genomic sequencing (9–12). Recently, it has been shown that variant prevalence in wastewater correlates with clinical data (13). Therefore variant monitoring in wastewater may serve as an efficient and complementary approach to genomic epidemiology based on individual patient samples.

However, it is challenging to analyze wastewater samples for their SARS-CoV-2 genomic composition, because concentrations of SARS-CoV-2 are low, samples are enriched in PCR inhibitors, viral genomes may be fragmented, and sewage contains large amounts of bacterial, human and other viral DNA and RNA genomes. In addition, the data quality obtained from sequencing the mixture of viral genomes is affected by amplification biases, sequencing errors, and incomplete phasing information, which further complicates the detection of an emerging viral lineage that is present only in a small fraction of infected persons.

Here, we address some of the key challenges and demonstrate that genomic sequencing of wastewater samples can be used for early detection, quantitative monitoring, and estimation of transmission fitness of any genetic variant of SARS-CoV-2.

## Methods

### Study overview

We collected a total of 122 samples from three Swiss wastewater treatment plants (WWTPs) located in Zurich, Lausanne and an alpine ski resort between July 2020 and February 2021 (Supplementary Figure 1A). These samples include a close-meshed time-series for Zurich and Lausanne between December 2020 and mid-February 2021. Viral RNA was extracted from raw influent samples and subjected to amplicon-based next-generation sequencing (NGS) using 2×250 bp paired-end sequencing (Figure 1A). We compared normalized amplicon coverage to clinical SARS-CoV-2 sequencing data to assess general data quality and performed replicate and spike-in experiments to assess reproducibility and quantifiability of SARS-CoV-2 variants in wastewater. Then, we searched the 122 wastewater samples for evidence of the respective signature mutations of the variants B.1.1.7, B.1.351, and P.1 (Supplementary Table S1). For early detection in individual samples, we developed a bioinformatics tool that searches for groups of signature mutations that can be observed directly on the same sequencing read pair originating from the same amplicon. We used the dense time-series data available for December 2020 to mid-February 2021 to estimate the local prevalence of the B.1.1.7 variant, calculated its fitness advantage and compared our results to estimates based on clinical data from the same geographical areas.

**Figure 1.**
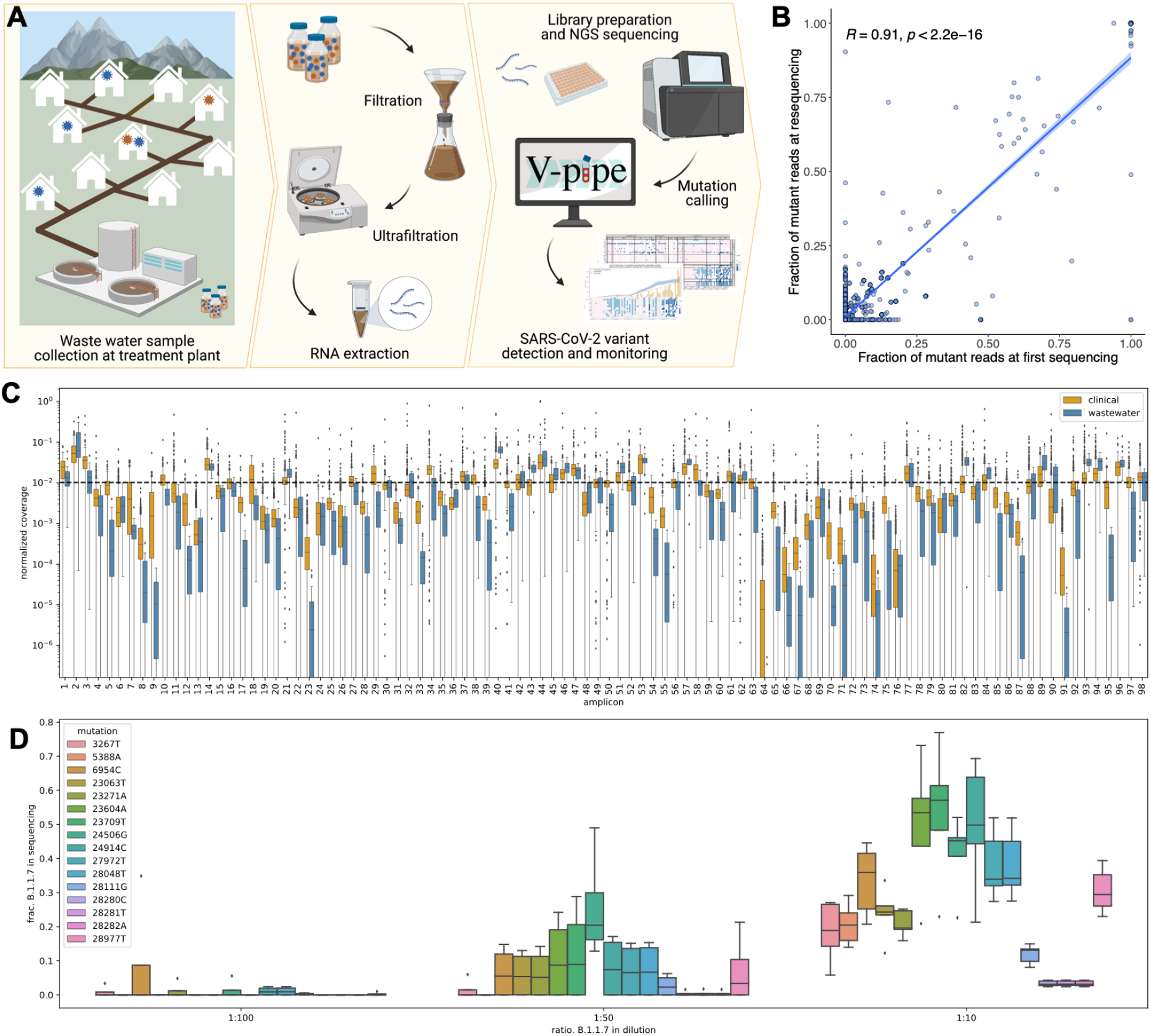
(A) Overview of the wastewater sampling campaign: (Left) Collection of raw wastewater samples containing a mixture of wild type and variant SARS-CoV-2 viral RNA. (Middle) Viral concentration and nucleic acid extraction. (Right) Amplification using ARTIC v3 primers, library preparation, next-generation sequencing (NGS), and mutation calling using V-Pipe is followed by statistical analysis to detect and quantify the presence of SARS-CoV-2 variants and estimate epidemiological parameters. (B) Reproducibility of B.1.1.7 prevalence based on resequencing of 25 samples. Each dot shows the fraction of B.1.1.7-compatible reads in two sequencing runs per sample and per signature mutation. Pearson correlation coefficient, R, indicates a high degree of reproducibility. (C) Per-amplicon normalized coverage distributions after quality filtering and alignment in the same NGS batch containing both clinical (orange) and wastewater (blue) samples. Per-amplicon absolute coverages can be found in Supplementary Figure S2. (D) Reproducibility of B.1.1.7 prevalence in a dilution series experiment. Boxplots represent fractions of substitutions called in five replicates of wastewater spiked with SARS-CoV-2 RNA at 3 different B.1.1.7-to-wild-type ratios.

### Wastewater sample collection and preparation

Raw wastewater samples were collected from three Swiss WWTPs: Werdhölzli, Zurich (64 samples, Jul 2020-Feb 2021, population connected: 450,000), Vidy, Lausanne (49 samples, Sep 2020-Feb 2021, population connected: 240,000), and an alpine ski resort (8 samples, Dec 2020) (Figure 1A, Supplementary Figure S1). Samples were concentrated and viral RNA was extracted as described before (14). In brief, 24-hour composite samples (Zurich and Lausanne) or grab samples (ski resort) were collected in 500 ml polystyrene or polypropylene plastic bottles, shipped on ice, and stored at 4°C for up to 8 days before processing. Aliquots of 50 ml were clarified by filtration (2 μm glass fiber filter (Millipore) followed by a 0.22 μm filter (Millipore), Zurich samples), or by centrifugation (4,863 xg for 30 minutes, Lausanne and ski resort samples). Clarified samples were then concentrated using centrifugal filter units (Centricon Plus-70 Ultrafilter, 10kDa, Millipore, USA) by centrifugation at 3,000 xg for 30 minutes. Centricon cups were inverted and the concentrate was collected by centrifugation at 1,000 xg for 3 minutes. The resulting concentrate (up to 280 μL) was extracted using the QiaAmp Viral RNA MiniKit (Qiagen, USA) according to the manufacturer’s instructions, adapted to the larger volumes, and eluted in 80 μL. Samples collected after February 1 were further purified using OneStep PCR Inhibitor Removal columns (Zymo Research, USA). RNA extracts were stored at -80°C for up to 4 months before sequencing.

### Genomic sequencing

RNA extracts from wastewater samples were used to produce amplicons and to prepare libraries according to the COVID-19 ARTIC v3 protocol(15) with minor modifications. Briefly, extracted RNA was reversed transcribed using the NEB LunaScript RT SuperMix Kit (New England Biolabs, USA) and the resulting cDNA was amplified with the ARTIC v3 panel from IDT(IDT, USA). The amplicons were end-repaired and polyadenylated before ligation of adapters using NEB Ultra II (New England Biolabs, USA). Fragments containing adapters on both ends were selectively enriched and barcoded with unique dual indexing with PCR. Libraries were sequenced using the Illumina NovaSeq 6000 and MiSeq platforms, resulting in paired-end reads of length 250 bp each.

### Mutation calling

NGS data was analyzed using V-pipe(16), a bioinformatics pipeline for end-to-end analysis of viral sequencing reads obtained from mixed samples. Individual low-frequency mutations were called based on local haplotype reconstruction using ShoRAH (17). For detecting mutation co-occurrence, we developed a novel computational tool called Cojac (CoOccurrence adJusted Analysis and Calling). The ARTIC v3 protocol relies on tiled amplification, and some amplicons cover multiple positions mutated in a variant (Supplementary Table S1). As the samples are sequenced with paired end 250 bp reads, each 400 bp amplicon can be fully observed on the read pairs in close to all instances. Detecting multiple signature mutations on the same amplicon increases the confidence of mutation calls at very low variant read counts. This opens the possibility of earlier detection, while variant concentrations are still too low for reliable detection of individual mutations. Cojac takes the multiple read alignments (BAM files) and counts read pairs with variant-specific mutational patterns. Cojac can be configured to work with any tiled amplification scheme and to simultaneously search for multiple variants, each defined by a list of signature mutations. Cojac is freely available at https://github.com/cbg-ethz/cojac/ or as a bioconda package at https://bioconda.github.io/recipes/cojac/README.html.

### Statistical data analysis

For Zurich, we used the 55 sequencing experiments (excluding 1 failed) covering 46 dates ranging from December 8, 2020 to February 11, 2021. For Lausanne, we used the 52 sequencing experiments (excluding 4 failed) covering 43 dates ranging from December 8, 2020 to February 13, 2021. When a WWTP sample was sequenced multiple times, we fixed the empirical frequencies of the B.1.1.7 signature mutations for a given day by averaging their values between the different sequencing experiments. We only used non-synonymous substitutions for quantification. Frequencies of the B.1.1.7 signature substitutions in wastewater-derived NGS data were resampled with replacement and averaged per wastewater sample, before being smoothed across time by local regression using locally weighted scatterplot smoothing (lowess) with ⅓ bandwidth from the Python v3.7.7 library statsmodels v0.12.1(18). This process was repeated 1000 times to construct bootstrap estimates of the B.1.1.7 per-day frequency curves. The smoothed resampled values were used to compute point estimates by averaging the daily B.1.1.7 prevalence as well as confidence intervals as the empirical 2.5% and 97.5% quantiles. For the prevalence of B.1.1.7 in clinical samples, we used the whole-genome sequencing data comprising randomly selected SARS-CoV-2-positive samples provided by Viollier AG, as described previously(19). Daily cantonal relative abundances of variants were estimated as their empirical frequencies in sequenced samples. For each canton, the sequenced cases were resampled with replacement and aggregated into daily relative frequencies of B.1.1.7, which was then smoothed temporally using the same lowess smoother as mentioned above. This process was repeated 1000 times to construct bootstrap estimates of the B.1.1.7 daily cantonal relative prevalence, which were aggregated into point estimates and confidence intervals by the same method as described above.

### Estimation of epidemiological parameters

Following Chen et al.(19), we assume that the relative frequency *p*(*t*)of the B.1.1.7 variant in the population at time *t* follows a logistic growth with rate a and inflection point *t*_0_,

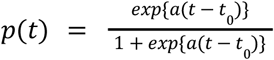

For the wastewater samples, we further assume that the B.1.1.7 signature mutation counts are distributed according to a binomial distribution, with expected value equal to *p*(*t*) times the total coverage at the respective site. Similarly, we assume that the B.1.1.7-positive clinical samples are also distributed according to a binomial distribution with expected value equal to *p*(*t*) times the number of clinical samples analyzed. The R v3.6.1 package stats(20) was used to produce maximum likelihood estimates of the model parameters with a generalized linear model. Confidence intervals were computed based on their asymptotically normal distribution. To account for overdispersion due to the inherently noisy nature of wastewater sequencing data, the confidence intervals were computed using the variance of a quasibinomial(21) distribution. Although clinical data are not expected to exhibit overdispersion, the same procedure was applied for the sake of consistency. Confidence bands were first generated for the linear predictors, and then back-transformed into confidence bands for the regression curves to ensure that they are restricted to the interval [0,1]. Estimates of the logistic growth parameter a were then transformed into estimates of the transmission fitness advantage *f*_*d*_ assuming the discrete-time model of Chen et al.(19) with generation time *g* = 4. 8 days, such that *f*_*d*_ = *exp*(*ag*) - 1. Confidence intervals for the logistic growth parameter awere then back-transformed into confidence intervals for the fitness advantage *f*_*d*_. This inference procedure was repeated at multiple timepoints with only the clinical and wastewater sequencing data available at these timepoints, to generate online estimates and confidence intervals of what could have been inferred about *f*_*d*_ at that time. These estimates were compared to the estimates of *f*_*d*_ reported in Chen et al.(19) for the Lake Geneva region (population 1.6 Mio), which includes Lausanne, and the Greater Zurich Area (population 1.5 Mio). The confidence intervals for these regional estimates of *f*_*d*_ were recomputed using back-transformation of the confidence intervals reported for the regional estimates of *a*, so that they could be meaningfully compared with the ones based on our data.

### Dilution experiment

RNA samples of cultivated SARS-CoV-2 wild type (Wuhan strain) and of a clinical B.1.1.7 strain were obtained. Each RNA sample was diluted in an RNA extract produced from SARS-CoV-2-free wastewater (November 2019, Lausanne) to a final concentration of 200 gc/μL. Wild type and B.1.1.7 solutions were then mixed at ratios of 10:1, 50:1 and 100:1, and each mixture was sequenced five times.

### Replicate experiment

RNA extract was produced as described above from two samples obtained from the Lausanne WWTP on January 7, 2020. The extracts were pooled and subsequently divided into 9 replicate samples for sequencing.

### Patient sequences

Per-patient SARS-CoV-2 consensus sequences were downloaded from GISAID(22) for all samples collected in Switzerland between February 24, 2020, and February 13, 2021, and not identified as either B.1.1.7, P.1, or B.1.351 (see Supplementary Material for the list of accession numbers).

## Results

We first assessed the quality of genomic sequencing data derived from wastewater samples. We found that the normalized amplicon coverage obtained from the wastewater samples was not significantly different from the coverage of clinical samples (Figure 1C) and that it allowed for calling low-frequency mutations in most genomic regions of most wastewater samples we analyzed (Supplementary Figure S2). The additional replicate and spike-in experiments indicate that the relative prevalence of genomic variants can be quantified from the NGS data and that replication increases precision, especially in the monitoring of low-frequency variants (Figures 1B,D, Supplementary Material).

The variant frequencies in the 122 wastewater samples revealed a continuous increase in the prevalence of the B.1.1.7 variant in Zurich starting around mid-December and in Lausanne starting in late December (Figure 2A). Much of the noise in the data can be removed by computing smoothed estimates over time and over signature mutations (Methods). When comparing these estimates of B.1.1.7 prevalence to those obtained from clinical samples, we found that they aligned very closely, even though the treatment plants serve only a subset of the respective cantonal populations (Zurich: 29% of canton Zurich, Lausanne: 30% of canton Vaud) (Figure 2B,C). For the alpine ski resort, we detected the B.1.1.7 variant over the whole period of observation (20 Dec 2020 - 29 Dec 2020), consistent with the popularity of the ski resort as a holiday destination with British tourists (Figure 2A). Unlike for B.1.1.7, we found almost no evidence for the distinctive signature mutations of B.1.351 or P.1 (Supplementary Material).

**Figure 2.**
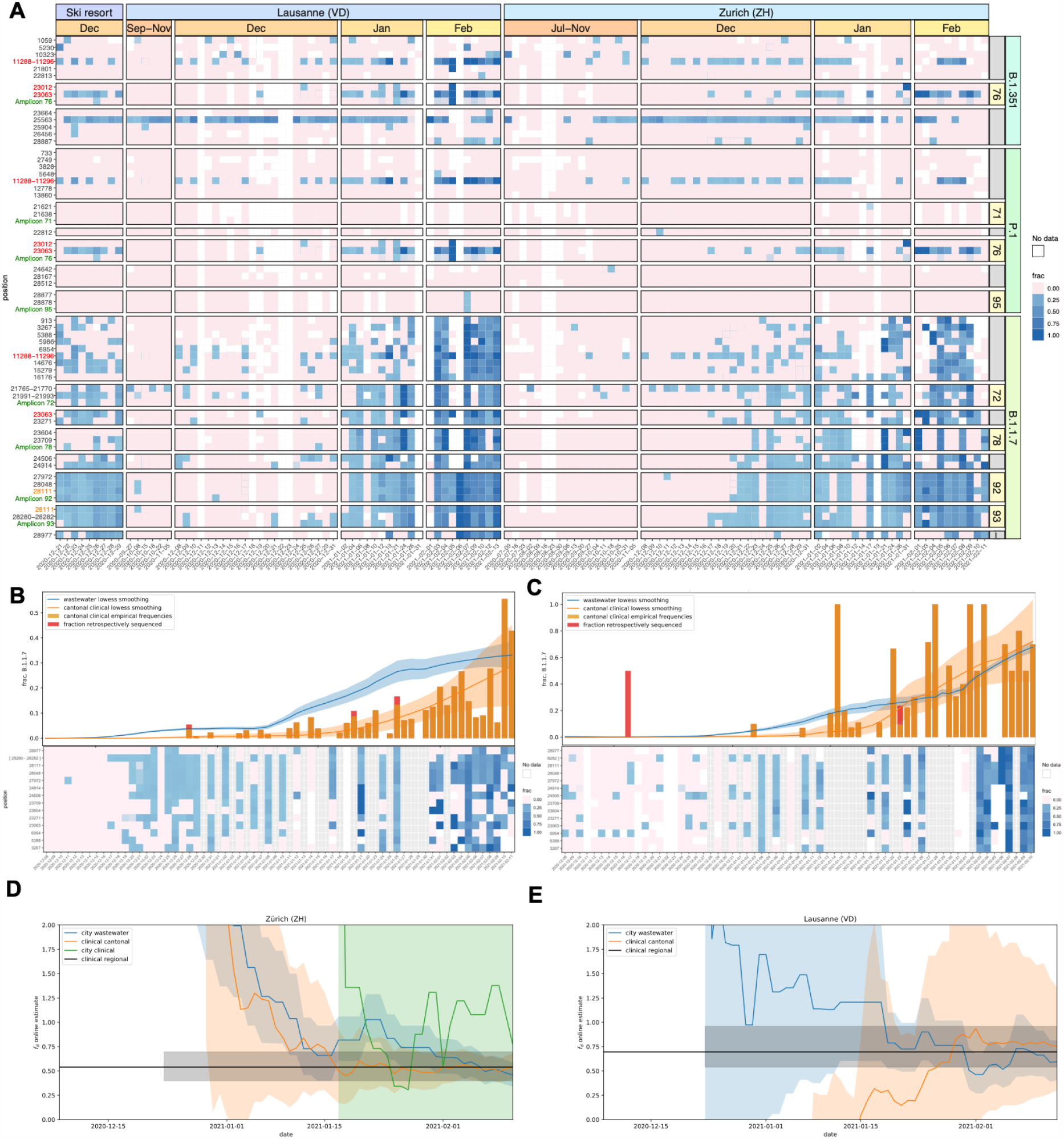
(A) Longitudinal surveillance of B.1.351, B.1.1.7, and P.1 signature mutations in wastewater samples collected at three Swiss WWTPs. Blue color shading encodes the observed fraction of each signature mutation in each sample, pink indicates absence of the mutation, and white indicates missing values (due to lack of sufficient coverage). Mutations are grouped by variant and further by amplicon number in case multiple mutations co-occur on the same amplicon. Rows labelled “Amplicon” followed by a number (green) show the observed frequency of co-occurrence on the same read pair for all mutations located on the respective amplicon. Mutations occur multiple times on the y-axis if they either occur in more than one variant (red) or are located on two overlapping amplicons (orange). (B, C) B.1.1.7 prevalence estimates based on wastewater sequencing data and on cantonal clinical sequencing data for Zurich (B) and Lausanne (C). Bottom panels show frequencies of B.1.1.7-characteristic substitutions found in wastewater sequencing samples, which are aggregated and smoothed in the top panel. Grey columns show dates without wastewater samples. Orange and red bars indicate the frequency of B.1.1.7-positive cantonal clinical samples, which are also smoothed. The red parts indicate fraction of B.1.1.7-positive samples that were sequenced retrospectively in March/April 2021 (cut-off date for the GISAID submission date, March 21 2021). Shaded areas represent 95% confidence bands. (D, E) Estimates of the transmission fitness advantage *f*_*d*_, computed online (see Methods) using the wastewater (blue), cantonal clinical (orange), and city clinical (green, Zurich only) sequencing data only until the respective timepoints for Zurich (D) and Lausanne (E). Shaded areas represent 95% confidence intervals. Horizontal black lines indicate offline estimates of *f*_*d*_ based on clinical samples of the larger geographical regions of Lake Geneva Region and Greater Zurich Area dated 14 Dec 2020 to 11 Feb 2021 from Chen et al.(19).

For early detection in individual samples, we additionally searched for co-occurrence of mutations on read pairs, which provides high confidence in the presence of the respective strain, as independent biological generation or technical artifacts are both very unlikely to produce such mutational patterns (Supplementary Material). We analyzed four amplicons that each contain two or three B.1.1.7-defining mutations, one amplicon with two signature mutations shared between B.1.351 and P.1, and one amplicon with two P.1 signature mutations. We found several co-occurrences (Figure 2A, Supplementary Table 3). The two mutations co-located on amplicon 93 provide the earliest evidence for B.1.1.7 in wastewater samples in Zurich on December 17 and in Lausanne on December 9. In both locations, these dates fall in a time when evidence based on single mutations alone was still very spotty (Figure 2A).

We compared our results to early variant detection based on clinical sequencing data in the respective cantons of Zurich and Vaud. In Switzerland around 4% of all SARS-CoV-2-positive clinical samples of December 2020 were sequenced. The first clinical evidence of the B.1.1.7 variant in canton Zurich was detected in a sample dated December 18, and in canton Vaud in a sample dated December 21. The former being one day later, the latter 13 days later than the first wastewater-based evidence. This finding is consistent with those of a retrospective sequencing campaign of March/April 2021 analyzing clinical samples collected in November and December 2020. The retrospectively obtained data revealed isolated occurrences of B.1.1.7 already on November 9 (3 cases) in canton Zurich, and on December 17 in canton Vaud (1 case) (Supplementary Table S5). For canton Vaud, that variant detection was still significantly later (8 days) than the first wastewater-based evidence in its capital Lausanne. For canton Zurich, the B.1.1.7-positive samples of November 9 all originated from municipalities located outside the catchment area of the studied treatment plant and therefore could not be detected in the analysed wastewater samples.

The co-occurring mutation pair on amplicon 93 we used for early detection in wastewater is highly specific to B.1.1.7, as it has only been observed 14 times (0.07%) outside of B.1.1.7 lineage in the 21,163 Swiss samples in the GISAID database (Supplementary Table S4) and only 138 times (0.01%) in all 1,397,333 SARS-CoV-2 GISAID samples until February 13, 2021. At later time points, we also observe evidence based on the other three B.1.1.7-specific amplicons both in Lausanne and Zurich. In the ski resort, evidence for the presence of B.1.1.7 has already been strong based on the analysis of individual mutations over the whole period of observation (December 20-29), and the amplicon-based analysis further supports this observation. For B.1.351 and P.1, the evidence based on co-occurrence is similarly weak as for the analysis of individual mutations (Supplementary Material). This result aligns with the clinical data for the period of study, with only one P.1 sample detected in canton Zurich (first occurrence on January 27) and none in canton Vaud. B.1.351 was detected two times in canton Zurich (first occurrence Jan 18) and seven times in canton Vaud (first occurrence Jan 14).

Next, we estimated the transmission fitness advantage of B.1.1.7 from its prevalence in wastewater for both Lausanne and Zurich (Methods). This parameter informs about the epidemiological relevance of an emerging variant. Our estimates of the transmission fitness advantage of 46% (CI 35%-60%) for the Zurich WWTP catchment and 59% (CI 42%-84%) for the Lausanne WWTP catchment are in line with those based on regional clinical data(19) and with reports from the United Kingdom(23) (Supplementary Material). Narrowing the clinical data down to the canton level, the estimates are still in line with the wastewater for Zurich (54%, CI 43%-69%, based on 2062 samples), while for Vaud the clinical estimates become less precise (75%, CI 34%-144%, based on 345 samples) reflected by the huge confidence interval. To assess how early the transmission fitness advantage can be estimated with acceptable precision, we also computed online estimates of the transmission fitness advantage, i.e., we used only the data up to the respective time point, 46 wastewater samples at most per location (Methods). For canton Vaud, the wastewater-based estimates for the Lausanne plant are more precise than the estimates based on hundreds of clinical samples from the canton (Figure 2D). For canton Zurich, the estimates are similar to those of thousands of cantonal clinical samples with one outlier around mid-January for Zurich (Figure 2E, Supplementary Material). Restricting the clinical data to the 115 samples from the city of Zurich which comprises the majority of the catchment area of the treatment plant, the precision of the wastewater-based online estimates are clearly superior (Figures 2D, Supplementary Material).

## Discussion

We have demonstrated how genomic sequencing of wastewater samples can be used to detect, monitor, and evaluate genetic variants of SARS-CoV-2 on a population level. Specifically, we have shown that the detection of the local outbreak of the B.1.1.7 variant in two Swiss cities was possible in wastewater before it was observed in clinical samples. This finding is especially remarkable in a country like Switzerland which has a high sequencing rate of SARS-CoV-2-positive clinical samples (4% in December 2020).

However, we also found that early variant detection based on single wastewater samples is very challenging, as initially only a small subset of signature mutations could be observed which makes it difficult to distinguish between signal and noise in the sequencing data. The high noise level in the data is attributable to the technical challenges that arise in the collection and processing of the raw wastewater sample, the extraction of SARS-CoV-2 RNA and its amplification. Our results suggest that high sampling density across time and replicate sequencing are key elements to improve the signal-to-noise ratio.

We also developed an approach to boost the signal strength in individual samples. It is based on the detection of co-occurring signature mutations on the same read pair, as their presence in a sample constitutes a much stronger signal than individual mutations. This approach was particularly valuable for the detection of B.1.1.7 which has multiple highly specific mutation pairs and even one triplet that can be detected in this manner. In Lausanne, the first co-occurrence-based evidence of the B.1.1.7 variant occurred 13 days prior to the first clinical evidence in the area. Among the other variants we studied, namely B.1.351 and P.1, only P.1 had one variant-specific mutation pair and one pair was shared among the two variants. The more recent variant of concern B.1.617.2 has two distinct co-occurrences that allow for distinguishing it from other current variants, including the other sub-variants of the B.1.617.^*^ lineage. No such co-occurrence can be found for the C.36.3 variant with the current amplicon tiling, but the redesigned amplicon tiling of the upcoming ARTIC v4 protocol (https://artic.network/) will provide one amplicon with three C.36.3-specific mutation sites. In general, the usefulness of the co-occurrence analysis for variant identification depends on the exclusivity of co-occurrences and is negatively affected by the presence of recurring mutations in separate lineages (24,25). As more and more variants arise, it may however become necessary to disentangle their aggregate signal in wastewater.

Besides early detection, we have shown that sequencing data obtained from wastewater samples can also be used to monitor the local prevalence of a variant and to estimate its speed of spreading earlier and based on substantially fewer samples as compared to using clinical samples. Moreover, wastewater samples have the advantage that undiagnosed asymptomatic cases are represented in the data, which are systematically overlooked in clinical sequencing. Therefore, genomic analysis of SARS-CoV-2 variants in wastewater samples can inform epidemiological studies and complement established approaches based on clinical samples.

All these findings suggest that the sequencing of wastewater samples is a highly valuable endeavor and that our methodology provides a blueprint for rapid, unbiased, and cost-efficient genomic surveillance of any SARS-CoV-2 variant based on repeated sequencing of wastewater samples.

## Supporting information

GISAID Acknowledgement Table

## Data Availability

A version of the wastewater sequencing data with the human reads removed is available on the Sequence Read Archive (SRA) under project accession number PRJEB44932

## Author Contributions

Conceptualization: NB, TK, TRJ, CO, TS, XFC, IT

Data curation: IT, CC, KPJ, SN

Formal Analysis: KJ, LF, DD

Funding acquisition: NB, TK, TRJ, CO, TS, XFC

Investigation: XFC, AK, PG, ES, CBä, CA, FC, LC, ATC

Methodology: NB, XFC, KJ, DD, IT

Project administration: NB, TK, TRJ, CO, TS

Software: IT, KPJ, LF, DD, KJ

Supervision: NB, TK, TRJ, CO, TS, IT

Validation: NB, IT, KJ, KPJ

Visualization: KJ, DD, KPJ

Writing – original draft: NB, KJ, DD, IT

Writing – review & editing: NB, TK, TRJ, CO, TS, KJ, KPJ, LF, XFC, DD, CA, SN, CC, CBä, CBe.

## Acknowledgements

RNA samples of cultivated SARS-CoV-2 wild type (Wuhan strain) and of a clinical B.1.1.7 strain for the dilution experiment were kindly provided by Tobias Schindler (Swiss Tropical and Public Health Institute). Library preparation and sequencing were performed at the Functional Genomics Center Zurich, ETHZ. The Supplementary Material contains the detailed acknowledgements of the originating and submitting laboratories of the GISAID data.

