## Supplement for "Detection and surveillance of SARS-CoV-2 genomic variants in wastewater"

### Supplementary material for “Detection and surveillance of SARS-CoV-2 genomic variants in wastewater”

#### Overview of signature mutations of B.1.1.7, B.1.351, and P.1

**Supplementary Table S1.** Signature mutations of the B.1.1.7, B.1.351, and P.1 lineages, their amplicon location and frequencies among all 21,163 non-B.1.1.7, non-B.1.351, and non-P.1 consensus sequences available in GISAID and obtained from clinical samples in Switzerland until February 13, 2021.

| | Nucleotide | | | Variant | | | | GISAID | |
|---|---|---|---|---|---|---|---|---|---|
| **Gene** | **position** | **change** | **Amino acid change** | **B.1.1.7** | **B.1.351** | **P.1** | **Ampli-cons** | **counts** | **relative freq. (%)** |
| **ORF1ab** | **733** | T>C | syn | 0 | 0 | 1 | [3] | 0 | 0 |
| **ORF1ab** | **913** | C>T | syn | 1 | 0 | 0 | [3] | 20 | 0.1 |
| **ORF1ab** | **1059** | C>T | T265I | 0 | 1 | 0 | [4] | 324 | 1.53 |
| **ORF1ab** | **2749** | C>T | syn | 0 | 0 | 1 | [9] | 0 | 0.00 |
| **ORF1ab** | **3267** | C>T | T1001I | 1 | 0 | 0 | [11] | 48 | 0.22 |
| **ORF1ab** | **3828** | C>T | S1188L | 0 | 0 | 1 | [13] | 5 | 0.02 |
| **ORF1ab** | **5230** | G>T | K1655N | 0 | 1 | 0 | [17] | 77 | 0.36 |
| **ORF1ab** | **5388** | C>A | A1708D | 1 | 0 | 0 | [18] | 35 | 0.16 |
| **ORF1ab** | **5648** | A>C | K1795Q | 0 | 0 | 1 | [19] | 1 | 0.00 |
| **ORF1ab** | **5986** | C>T | syn | 1 | 0 | 0 | [20] | 57 | 0.27 |
| **ORF1ab** | **6954** | T>C | I2230T | 1 | 0 | 0 | [23] | 28 | 0.13 |
| **ORF1ab** | **10323** | A>G | K3353R | 0 | 1 | 0 | [34] | 261 | 1.23 |
| **ORF1ab** | **11288** | --------- | 3675-3677 -SGF | 1 | 1 | 1 | [37] | 150 | 0.71 |
| **ORF1ab** | **12778** | C>T | syn | 0 | 0 | 1 | [42, 43] | 2 | 0.01 |
| **ORF1ab** | **13860** | C>T | syn | 0 | 0 | 1 | [46] | 66 | 0.31 |
| **ORF1ab** | **14676** | C>T | syn | 1 | 0 | 0 | [49] | 32 | 0.15 |
| **ORF1ab** | **15279** | C>T | syn | 1 | 0 | 0 | [51] | 62 | 0.29 |
| **ORF1ab** | **16176** | T>C | syn | 1 | 0 | 0 | [53, 54] | 29 | 0.14 |
| **S** | **21621** | C>A | T20N | 0 | 0 | 1 | [71] | 0 | 0 |
| **S** | **21638** | C>T | P26S | 0 | 0 | 1 | [71] | 38 | 0.18 |
| **S** | **21765** | ------ | 69-70 -HV | 1 | 0 | 0 | [72] | 1054 | 4.98 |
| **S** | **21801** | A>C | D80A | 0 | 1 | 0 | [72] | 0 | 0.00 |
| **S** | **21991** | --- | 144 -Y | 1 | 0 | 0 | [72, 73] | 68 | 0.32 |
| **S** | **22812** | A>C | K417T | 0 | 0 | 1 | [75] | 0 | 0 |

| | | | | | | | | | |
|---|---|---|---|---|---|---|---|---|---|
| **S** | **22813** | G>T | K417N | 0 | 1 | 0 | [75] | 0 | 0.00 |
| **S** | **23012** | G>A | E484K | 0 | 1 | 1 | [76] | 43 | 0.20 |
| **S** | **23063** | A>T | N501Y | 1 | 1 | 1 | [76] | 68 | 0.32 |
| **S** | **23271** | C>A | A570D | 1 | 0 | 0 | [77] | 25 | 0.12 |
| **S** | **23604** | C>A | P681H | 1 | 0 | 0 | [78] | 243 | 1.15 |
| **S** | **23664** | G>T | A701V | 0 | 1 | 0 | [78] | 4 | 0.02 |
| **S** | **23709** | C>T | T716I | 1 | 0 | 0 | [78] | 123 | 0.58 |
| **S** | **24506** | T>G | S982A | 1 | 0 | 0 | [81] | 20 | 0.09 |
| **S** | **24642** | C>T | T1027I | 0 | 0 | 1 | [81] | 24 | 0.11 |
| **S** | **24914** | G>C | D1118H | 1 | 0 | 0 | [82] | 27 | 0.13 |
| **ORF3a** | **25563** | G>T | Q57H | 0 | 1 | 0 | [84] | 6532 | 30.87 |
| **ORF3a** | **25904** | C>T | S171L | 0 | 1 | 0 | [85] | 49 | 0.23 |
| **E** | **26456** | C>T | P71L | 0 | 1 | 0 | [87] | 16 | 0.08 |
| **ORF8** | **27972** | C>T | Q27stop | 1 | 0 | 0 | [92] | 62 | 0.29 |
| **ORF8** | **28048** | G>T | R52I | 1 | 0 | 0 | [92] | 62 | 0.29 |
| **ORF8** | **28111** | A>G | Y73C | 1 | 0 | 0 | [92, 93] | 23 | 0.11 |
| **ORF8** | **28167** | G>A | E92K | 0 | 0 | 1 | [93] | 7 | 0.03 |
| **N** | **28280** | GAT>CTA | D3L | 1 | 0 | 0 | [93] | 29 | 0.14 |
| **N** | **28512** | C>G | P80R | 0 | 0 | 1 | [94] | 0 | 0 |
| **N** | **28887** | C>T | T205I | 0 | 1 | 0 | [95] | 178 | 0.84 |
| **N** | **28977** | C>T | S235F | 1 | 0 | 0 | [95] | 42 | 0.2 |

#### Sampling locations and sample overview

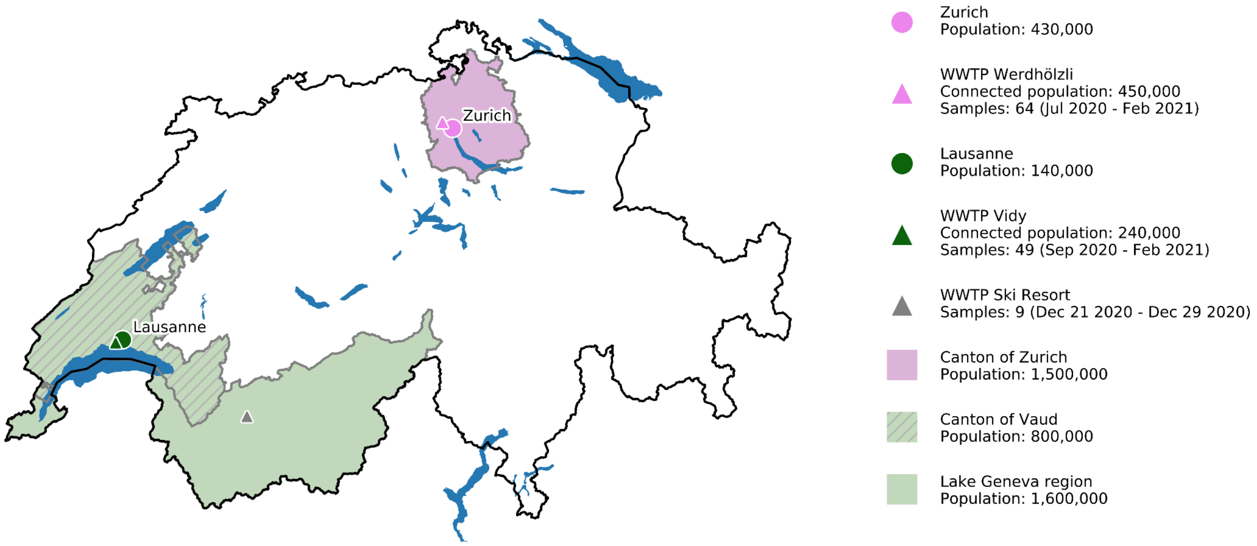


**Supplementary Figure S1.** Geographical locations of wastewater treatment plants (WWTPs) and sample information. WWTP Werdhölzli is located in the canton Zurich and serves the city of Zurich and part of the canton. WWTP Vidy is located in the canton Vaud and serves the city of Lausanne and part of the canton. The canton Vaud is part of the Lake Geneva Region from which clinical samples are analyzed in Figure 2E (clinical regional samples).

#### Quality of NGS data

DNA libraries were prepared from the viral RNA extracted from the wastewater samples using the ARTIC V3 protocol with minor modifications. Briefly, extracted RNA was reversed transcribed using the NEB LunaScript RT SuperMix Kit (E3010L) and the resulting cDNA was amplified with the ARTIC v3 panel from IDT(10006788). The amplicons were end-repaired and polyadenylated before ligation of adapters using NEB Ultra II (E7645L). Fragments containing adapters on both ends were selectively enriched and barcoded with unique dual indexing with PCR. Libraries were sequenced using 2 x 250bp paired-end reads (Methods). After quality control, we obtained a median number of reads per sample of 949,884. In the subsequent read mapping step, we successfully aligned a median of 472,278 reads per sample, which resulted in a median per-sample median coverage of 1539 reads per position of the SARS-CoV-2 genome (range 0 - 28,958).

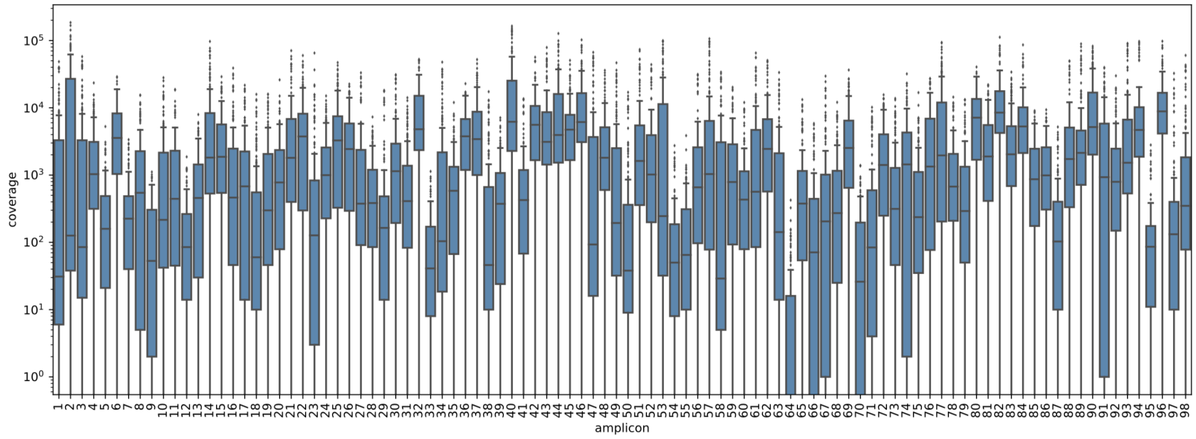


**Supplementary Figure S2.** Distributions of per-amplicon coverage in all 122 wastewater samples after quality filtering and alignment.

#### Reproducibility of variant detection and quantification

We selected 24 samples (Supplementary Table 2) for resequencing in a separate sequencing run and compared the position- and sample-specific fraction of reads supporting B.1.1.7 signature mutations (Figure 1B). Most of the resequenced samples originate from the second half of December 2020 to early January 2021, the time frame in which the B.1.1.7 variant emerged in the Swiss population. We observe a high correlation between the fraction pairs (Pearson correlation coefficient $R^2$ = 0.83, $p < 2.2\times10^{-16}$) which indicates a high degree of reproducibility.

To assess the influence of variant concentration on sensitivity, we sequenced pre-pandemic wastewater samples spiked with mixes of SARS-CoV-2 RNA at three different B.1.1.7-to-wild-type ratios, 10:1, 50:1 and 100:1 (Methods). Each mixture was sequenced five times (Fig 1D). Overall, the estimated fraction of B.1.1.7 displayed high correlation between replicates: in an analysis of variance (ANOVA) dilution ratios accounted for 45% of the variance in B.1.1.7 mutation frequencies, with higher relative precision at higher concentration (Supplementary Figure S3). Mutation positions accounted for 20% of the variance in B.1.1.7 mutation frequencies (while controlling for dilution), with some mutation frequencies consistently underestimated or overestimated, possibly due to background prevalence of mutations or effects of mutations on primer annealing.

**Supplementary Table S2.** List of 25 samples resequenced to assess reproducibility of B.1.1.7 prevalence estimates (Fig. 1B). Samples have been sequenced two times in total, except for the sample taken in Lausanne on October 15 which was sequenced three times.

| Sample | Date | Location |
|---|---|---|
| C6_12_2021_01_04 | 2021-01-04 | Lausanne (VD) |
| G2_12_2021_01_02 | 2021-01-02 | Lausanne (VD) |
| F2_12_2020_12_31 | 2020-12-31 | Lausanne (VD) |
| E2_12_2020_12_27 | 2020-12-27 | Lausanne (VD) |
| D2_12_2020_12_25 | 2020-12-25 | Lausanne (VD) |
| C2_12_2020_12_23 | 2020-12-23 | Lausanne (VD) |
| B2_12_2020_12_19 | 2020-12-19 | Lausanne (VD) |
| G1_12_2020_12_17 | 2020-12-17 | Lausanne (VD) |
| A2_12_2020_12_15 | 2020-12-15 | Lausanne (VD) |
| F1_12_2020_12_13 | 2020-12-13 | Lausanne (VD) |
| E1_12_2020_12_10 | 2020-12-10 | Lausanne (VD) |
| D1_12_2020_12_09 | 2020-12-09 | Lausanne (VD) |
| C1_12_2020_12_08 | 2020-12-08 | Lausanne (VD) |
| A1_12_2020_10_15 | 2020-10-15 | Lausanne (VD) |
| B1_12_2020_10_15_n | 2020-10-15 | Lausanne (VD) |
| D4_10_2021_01_04 | 2021-01-04 | Zurich (ZH) |
| C4_10_2021_01_02 | 2021-01-02 | Zurich (ZH) |
| B4_10_2020_12_31 | 2020-12-31 | Zurich (ZH) |
| A4_10_2020_12_29 | 2020-12-29 | Zurich (ZH) |
| H3_10_2020_12_25 | 2020-12-25 | Zurich (ZH) |
| G3_10_2020_12_23 | 2020-12-23 | Zurich (ZH) |
| F3_10_2020_12_21 | 2020-12-21 | Zurich (ZH) |
| H1_10_2020_12_20 | 2020-12-20 | Zurich (ZH) |
| E3_10_2020_12_19 | 2020-12-19 | Zurich (ZH) |
| D3_10_2020_12_17 | 2020-12-17 | Zurich (ZH) |

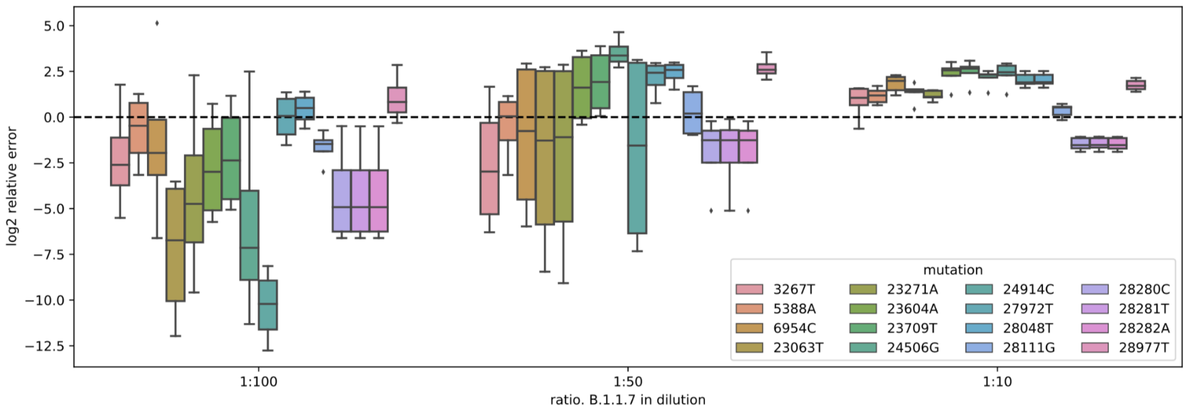


**Supplementary Figure S3.** Estimation error in dilution experiments. Log2 of relative overestimation/underestimation of the fraction of B.1.1.7 characteristic substitutions are reported, i.e. Log2 of proportion of mutation in sequencing data minus log2 of the proportion of B.1.1.7 RNA in dilution, for different substitutions at different dilution ratios.

To further assess reproducibility, RNA extracts from two samples from the Lausanne WWTP of January 7, 2021 were pooled and subsequently divided into 9 replicate samples for sequencing. B.1.1.7 fractions were estimated independently for each replicate. Estimates ranged from 0 to 0.1, had an average of 0.04 and a variance of 0.002 (Supplementary Figure S4), while the estimate of the prevalence of B.1.1.7 from temporally smoothed Lausanne WWTP samples was 0.07 (Figure 2C). In the seven replicate samples with evidence of B.1.1.7, we detected a median of five out of 16 B.1.1.7 characteristic nonsynonymous substitutions.

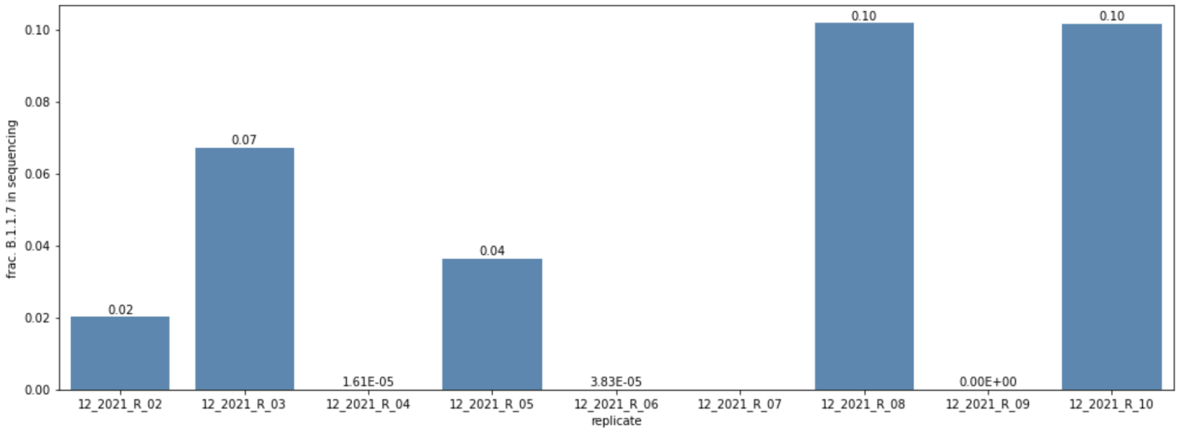


**Supplementary Figure S4.** Estimates of the fraction of B.1.1.7 in the resequencing experiments of the January 7, 2021, Lausanne WWTP sample. Estimate from the smoothed time series is ~0.07.

We take these results to indicate that quantification of the prevalence of a variant is possible, and that replication can increase the precision, especially when monitoring low frequency variants.

#### Detection of B.1.351 and P.1 in the wastewater samples

Unlike for B.1.1.7, we found almost no evidence for the distinctive signature mutations of B.1.351 or P.1. The frequently observed mutations G25563T (B.1.351) and A23063T (B.1.351 and P.1) are non-distinctive, as the former has been observed independently in the Swiss population prior to the emergence of B.1.351 and the latter is shared with the more prevalent B.1.1.7 variant. The absence of B.1.351 and P.1 in our data is consistent with the extremely low prevalence of these variants in Switzerland estimated from clinical samples. Between January and February 2021, out of 1338 clinical samples sequenced in Zurich and 466 in Vaud, P.1 was detected only once in Zurich and not at all in Vaud, while B.1.351 was observed two times in Zurich and seven times in Vaud.

#### Detection of mutation co-occurrence using Cojac

To validate Cojac, we first searched for the characteristic mutational patterns in two B.1.1.7-positive, two B.1.351-positive, and one P.1-positive control patient samples and indeed detected co-occurrence of the respective mutations on essentially all reads (Supplementary Table 3, last five rows). In addition, all patient and wastewater samples displayed, as expected, on amplicon 77 the A23403G signature mutation of the B.1 lineage, which is the most prevalent lineage (>99%) in Switzerland at the time of sampling[1,2].

We then analysed all 122 wastewater samples from the time series in Fig. 2A with cojac. Supplementary Table 2 shows the results for a subset of samples. The full cojac output can be found in Supplementary File 1. For Lausanne, first evidence of the B.1.1.7 variant appears on December 9 in the form of the two co-occurring signature mutations on amplicon 93. This is around the same time as we start to detect individual B.1.1.7 signature mutations in Lausanne. GISAID based background frequencies of these two mutations are extremely low (Supplementary Table 4). The combination of both mutations has only been observed in 0.07% of the 21,163 Swiss clinical samples that were not classified as B.1.1.7, B.1.351, or P.1 (until February 13, 2021). This makes it highly unlikely that the observed signal is originating from other sources than B.1.1.7-infected individuals. Single amplicon-based co-occurrence of B.1.1.7 signature mutations continues throughout December (Supplementary File 1). In early January, we start to see direct evidence of co-occurrence in three and later all four amplicons. (Supplementary Table 3, Supplementary File 1).

Co-occurrence of N501Y and E484K which are shared by B.1.351 and P.1 is first observed on January 21 (amplicon 76). On February 5, all 135 read pairs covering the amplicon exhibit the two mutations, while two days later, we find hardly any evidence of the co-occurrence. This volatility in combination with the fact that a subset of the B.1.1.7 strain has also acquired the E484K mutation in addition to the pre-existing N501Y mutation makes this particular co-occurrence a suboptimal marker for B.1.351 and P.1 detection. For P.1, two mutations occurring on amplicon 95 provide another opportunity for detecting the variant. However, so far only a single sample (February 7) provides evidence of the co-occurrence of these mutations. Looking at background mutation frequencies, we also observe that this mutation pair is not exclusive to P.1, but has been observed in 47 out of the 21,163 Swiss GISAID samples not classified as P.1, B.1.1.7, or B.1.351. Therefore, also the amplicon-based analysis for Lausanne does not provide clear evidence for the spread of B.1.351 or P.1.

For Zurich, the first weak evidence of the B.1.1.7 variant appears on December 17 in the form of the two co-occurring mutations on amplicon 93 which was also the earliest indicator of the variant in Lausanne. As discussed above, these mutations are unlikely to appear due to other sources than B.1.1.7-infected individuals. Like for Lausanne, we later also observe mutation co-occurrence based on the other three B.1.1.7-specific amplicons. Three out of four are observed for the first time on December 22, all four on December 25. The samples from Zurich analysed in this study did not provide any evidence for amplicon-based co-occurrence of B.1.351- or P.1-specific mutations.

Finally, the daily samples of the ski resort for the time interval between December 20 and 29 consistently show amplicon-based co-occurrence of B.1.1.7 mutations, with between one and three amplicons carrying the respective combinations of mutations.

**Supplementary Table S3.** Co-occurrence of signature mutations. For a subset of selected samples, genomic regions containing two or more B.1.1.7, B.1.351, or P.1 signature mutations observable on individual read pairs were analyzed. The first column indicates the sample date and location, or ID. Columns 2-7 are labeled with amplicons, their respective genomic regions and the signature mutations they contain. Displayed in columns 2-7 are the number of amplicons with all signature mutations present simultaneously, (separated by a slash) the total coverage by single amplicons, and the fraction of co-occurring mutant reads. Column seven reports the signature mutation (D614G) of lineage B.1, as a control. A variant is considered to be present in a sample if at least 5 read pairs and at least 0.1% of the total number of read pairs covering the amplicon carry all signature mutations of that region. The table shows a small subset of the wastewater samples and five patient samples known to derive from the B.1.1.7, B.1.351, and P.1 lineages as positive controls. Colors indicate observations indicating the presence of B.1.1.7 (yellow), B.1.351 or P.1 (red), P.1 (blue) and B.1 (purple). Results from samples that were sequenced twice appear in two separate rows.

| Sample | **Amplicon 72 21682-22013**<br><br>**21765-21770Δ, 21991-21993Δ (B.1.1.7)** | **Amplicon 78 23466-23822**<br><br>**C23604A, C23709T (B.1.1.7)** | **Amplicon 92 27809-28144**<br><br>**C27972T, G28048T, A28111G (B.1.1.7)** | **Amplicon 93 28105-28441**<br><br>**A28111G, 28280GAT→CTA (B.1.1.7)** | **Amplicon 76 22822-23188**<br><br>**G23012A, A23063T (B.1.351, P.1)** | **Amplicon 95 28699-29041**<br><br>**A28877T, G28878C (P.1)** | **Amplicon 77 23145-23499**<br><br>**A23403G (B.1)** |
|---|---|---|---|---|---|---|---|
| 2020-12-25 Ski-resort | 362 / 2729 13.26% | 0 / 1114 0.00% | 87 / 892 9.75% | 39 / 1143 3.41% | 0 / 2273 0.00% | 0 / 91 0.00% | 1379 / 1380 99.93% |
| 2020-12-23 Ski-resort | 257 / 1194 21.52% | 0 / 368 0.00% | 25 / 316 7.91% | 109 / 706 15.44% | 0 / 920 0.00% | 0 / 53 0.00% | 586 / 586 100.00% |
| 2020-12-21 Ski-resort | 0 / 990 0.00% | 0 / 2367 0.00% | 514 / 3689 13.93% | 0 / 20672 0.00% | 0 / 165 0.00% | 0 / 788 0.00% | 36208 / 36209 100.00% |
| 2021-02-07 Lausanne | 6177 / 9289 66.50% | 2393 / 2394 99.96% | 2887 / 4449 64.89% | 7004 / 7664 91.39% | 1 / 8556 0.01% | 20 / 435 4.60% | 6945 / 6947 99.97% |
| 2021-02-05 Lausanne | 0 / 13568 0.00% | 0 / 0 NA | 1492 / 4650 32.09% | 0 / 2211 0.00% | 135 / 135 100.00% | 0 / 133 0.00% | 0 / 0 NA |
| 2021-01-21 Lausanne | 662 / 6356 10.42% | 306 / 988 30.97% | 143 / 815 17.55% | 169 / 711 23.77% | 31 / 27238 0.11% | 0 / 4 0.00% | 586 / 586 100.00% |
| 2021-01-12 Lausanne | 16 / 128 12.50% | 6 / 30 20.00% | 2 / 5 40.00% | 30 / 354 8.47% | 3 / 26931 0.01% | 0 / 0 NA | 69 / 69 100.00% |
| 2021-01-04 Lausanne | 0 / 3112 0.00% | 105 / 3963 2.65% | 26 / 999 2.60% | 47 / 742 6.33% | 0 / 8232 0.00% | 0 / 376 0.00% | 621 / 623 99.68% |
| 2021-01-04 Lausanne | 0 / 2852 0.00% | 0 / 595 0.00% | 32 / 360 8.89% | 49 / 670 7.31% | 0 / 6909 0.00% | 0 / 27 0.00% | 825 / 826 99.88% |
| 2020-12-25 Lausanne | 0 / 524 0.00% | 0 / 448 0.00% | 0 / 47 0.00% | 21 / 219 9.59% | 0 / 16720 0.00% | 0 / 150 0.00% | 213 / 213 100.00% |
| 2020-12-21 Lausanne | 0 / 4 0.00% | 0 / 21 0.00% | 0 / 10 0.00% | 93 / 3393 2.74% | 0 / 0 NA | 0 / 2 0.00% | 10 / 10 100.00% |

| | | | | | | | |
|---|---|---|---|---|---|---|---|
| 2020-12-17 Lausanne | 0 / 460 0.00% | 0 / 322 0.00% | 5 / 457 1.09% | 0 / 40213 0.00% | 0 / 76 0.00% | 0 / 13 0.00% | 12846 / 12847 99.99% |
| 2020-12-09 Lausanne | 0 / 2338 0.00% | 0 / 1894 0.00% | 0 / 295 0.00% | 15 / 352 4.26% | 0 / 5092 0.00% | 0 / 13 0.00% | 276 / 276 100.00% |
| 2020-12-29 Zürich | 0 / 2776 0.00% | 0 / 1152 0.00% | 16 / 828 1.93% | 18 / 879 2.05% | 0 / 4093 0.00% | 0 / 97 0.00% | 1250 / 1250 100.00% |
| 2020-12-29 Zürich | 56 / 1906 2.94% | 54 / 1489 3.63% | 34 / 645 5.27% | 17 / 550 3.09% | 0 / 5084 0.00% | 0 / 17 0.00% | 410 / 410 100.00% |
| 2020-12-25 Zürich | 187 / 2689 6.95% | 0 / 1387 0.00% | 0 / 1331 0.00% | 15 / 1070 1.40% | 0 / 2303 0.00% | 0 / 178 0.00% | 1961 / 1961 100.00% |
| 2020-12-25 Zürich | 416 / 4034 10.31% | 345 / 4009 8.61% | 151 / 1627 9.28% | 152 / 1431 10.62% | 1 / 4270 0.02% | 0 / 325 0.00% | 712 / 730 97.53% |
| 2020-12-23 Zürich | 0 / 2023 0.00% | 0 / 953 0.00% | 32 / 957 3.34% | 1 / 1059 0.09% | 0 / 3245 0.00% | 0 / 158 0.00% | 1815 / 1815 100.00% |
| 2020-12-23 Zürich | 0 / 723 0.00% | 18 / 414 4.35% | 18 / 266 6.77% | 4 / 531 0.75% | 0 / 1327 0.00% | 0 / 89 0.00% | 75 / 75 100.00% |
| 2020-12-22 Zürich | 149 / 3453 4.32% | 0 / 946 0.00% | 30 / 1282 2.34% | 114 / 1642 6.94% | 0 / 5092 0.00% | 0 / 160 0.00% | 2581 / 2581 100.00% |
| 2020-12-20 Zürich | 0 / 6509 0.00% | 0 / 10734 0.00% | 154 / 13504 1.14% | 0 / 82020 0.00% | 0 / 802 0.00% | 0 / 1141 0.00% | 93625 / 93659 99.96% |
| 2020-12-20 Zürich | 0 / 652 0.00% | 0 / 428 0.00% | 5 / 89 5.62% | 4 / 295 1.36% | 0 / 5925 0.00% | 0 / 94 0.00% | 142 / 142 100.00% |
| 2020-12-17 Zürich | 0 / 5906 0.00% | 0 / 9112 0.00% | 0 / 9208 0.00% | 0 / 38732 0.00% | 0 / 233 0.00% | 0 / 887 0.00% | 34928 / 34939 99.97% |
| 2020-12-17 Zürich | 0 / 2459 0.00% | 0 / 994 0.00% | 0 / 639 0.00% | 5 / 859 0.58% | 0 / 9224 0.00% | 0 / 40 0.00% | 600 / 600 100.00% |
| Clinical B.1.351 sample 410256 | 0 / 8314 0.00% | 0 / 23023 0.00% | 0 / 20487 0.00% | 0 / 16822 0.00% | 156 / 156 100.00% | 0 / 23360 0.00% | 32633 / 32699 99.80% |
| Clinical B.1.351 sample 410279 | 0 / 1354 0.00% | 0 / 1401 0.00% | 0 / 2601 0.00% | 0 / 3526 0.00% | 8 / 8 100.00% | 0 / 3738 0.00% | 6570 / 6574 99.94% |
| Clinical B.1.1.7 sample 420389 | 212 / 214 99.07% | 236 / 236 100.00% | 389 / 389 100.00% | 1498 / 1501 99.80% | 0 / 3 0.00% | 0 / 418 0.00% | 3184 / 3184 100.00% |
| Clinical B.1.1.7 sample 420394 | 82 / 82 100.00% | 109 / 109 100.00% | 207 / 207 100.00% | 739 / 742 99.60% | 0 / 7 0.00% | 0 / 617 0.00% | 2067 / 2068 99.95% |
| Clinical P.1 sample 471206 | 0 / 2796 0.00% | 0 / 3634 0.00% | 0 / 47 0.00% | 0 / 1028 0.00% | 2660 / 2676 99.40% | 247 / 259 95.37% | 429 / 429 100.00% |

**Supplementary Table S4.** Background frequencies of mutation co-occurrence in genomic regions (amplicons) containing two or more B.1.1.7, B.1.351, or P.1 signature mutations. Frequencies are based on the 21,163 non-B.1.1.7, non-B.1.351, and non-P.1 consensus sequences available in GISAID and obtained from clinical samples in Switzerland until February 13, 2021.

| Variant | Amplicon | Mutations | Absolute frequency | Relative frequency (%) |
|---|---|---|---|---|
| B.1.1.7 | 72 | del 21765-21770 | 1054 | 4.98 |
| | | del 21991-21993 | 68 | 0.32 |
| | | co-occurrence | 16 | 0.08 |
| | 78 | C23604A | 243 | 1.15 |
| | | C23709T | 123 | 0.58 |
| | | co-occurrence | 35 | 0.17 |
| | 92 | C27972T | 62 | 0.29 |
| | | G28048T | 62 | 0.29 |
| | | A28111G | 23 | 0.11 |
| | | co-occurrence | 12 | 0.06 |
| | 93 | A28111G | 23 | 0.11 |
| | | GAT28280CTA | 29 | 0.14 |
| | | co-occurrence | 14 | 0.07 |
| B.1.351/ P.1 | 76 | G23012A | 43 | 0.20 |
| | | A23063T | 68 | 0.32 |
| | | co-occurrence | 0 | 0.00 |
| P.1 | 95 | A28877T | 50 | 0.24 |
| | | G28878C | 47 | 0.22 |
| | | co-occurrence | 47 | 0.22 |
| B.1 | 77 | A23403G | 20710 | 97.86 |

#### Estimation of epidemiological parameters

For any emerging variant, it is important to estimate its transmission fitness as early as possible in order to evaluate whether the variant may be of concern or not. To assess whether wastewater-based genomics can inform about fitness of the B.1.1.7 variant, we fit a discrete time logistic growth epidemiological model[3] to the increase in B.1.1.7 prevalence (Supplementary Figure S5). We estimated a transmission fitness advantage of 46% (CI 35%-60%) for Zurich and 59% (CI 42%-84%) for Lausanne. These estimates are coherent with those based on the regional clinical data reported in Chen et al. [3], namely 52% (40%-70%) for the greater Zurich area (based on 1590 samples dated December 14 2020 to February 11 2021) and 71% (54%-96%) for the Lake Geneva region around Lausanne (based on 548 samples dated December 14 2020 to February 11 2021). They are also in line with the 40%-70% transmission fitness advantage of B.1.1.7 that has been reported in the United Kingdom [4]. To examine how early the transmission fitness advantage could be meaningfully estimated, we fit the model multiple times on the portions of data available up to different timepoints, using data from wastewater, cantonal clinical samples, and city clinical data (Figure 2D,E). For Zurich, the estimates based on wastewater and cantonal clinical samples appear to follow the same trajectory, with the latter stabilizing about a week earlier. The delay in stabilization of the estimates derived from Zurich WWTP samples in comparison to those based on cantonal clinical data appears to be driven by two outliers in mid-January (Supplementary Figure S5). In contrast, the estimates based on the city-wide sequencing data (of the city having the greatest overlap in population with the population connected to the WWTP) are not informative during this timeframe. For Lausanne, the estimates based on wastewater appear to start converging weeks before those based on cantonal clinical samples. The Lausanne city (having the greatest overlap in population with the population connected to the Vidy WWTP) clinical data did not contain any B.1.1.7-positive sample. Thus, the online fitness estimates based on 46 and 43 pooled wastewater samples tend to be similar and in some cases superior to those obtained from 2062 and 345 individual clinical samples, respectively.

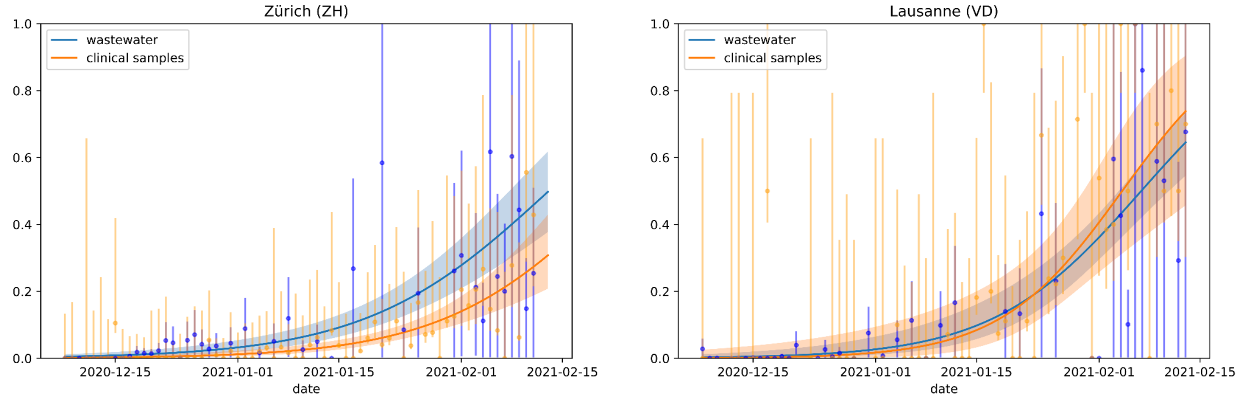


**Supplementary Figure S5.** Logistic growth model fitted to variant proportions derived from wastewater and clinical samples. Dots with error bars represent daily empirical proportions of B.1.1.7-positive clinical samples (orange), or average empirical proportions of B.1.1.7-characteristic substitutions in wastewater (blue). Error bars are 95% Wilson confidence intervals. Shaded areas represent 95% confidence bands for the logistic curve fits.

#### Early B.1.1.7-positive clinical samples

**Supplementary Table S5.** Overview of all early clinical samples from canton Zurich and canton Vaud that were identified as B.1.1.7 (cut-off date December 22, 2020). Grey rows correspond to samples that were only detected in a retrospective sequencing campaign conducted in March/April 2020 and therefore were not available for early detection of the B.1.1.7 variant.

| Sample | Sample date | Canton | Variant | Originating lab | Submitting lab | Submission date |
|---|---|---|---|---|---|---|
| Switzerland/VD-ETHZ-560572/2020 | Dec 17, 2020 | Vaud | B.1.1.7 | Viollier AG | D-BSSE, ETH Zurich | Apr 22, 2021 |
| Switzerland/VD-UZH-IMV144/2020 | Dec 21, 2020 | Vaud | B.1.1.7 | Dr. Boubaker Karim laboratory | Institute of Medical Virology, University of Zurich | Jan 5, 2021 |
| Switzerland/VD-ETHZ-6/2020 | Dec 21, 2020 | Vaud | B.1.1.7 | MCL Medizinische Laboratorien Hauptstandort Niederwangen | D-BSSE, ETH Zurich | Mar 3, 2021 |
| Switzerland/VD-UZH-IMV143/2020 | Dec 22, 2020 | Vaud | B.1.1.7 | Dr. Boubaker Karim laboratory | Institute of Medical Virology, University of Zurich | Jan 5, 2021 |
| Switzerland/ZH-ETHZ-500087/2020 | Nov 09, 2020 | Zurich | B.1.1.7 | Viollier AG | D-BSSE, ETH Zurich | Mar 2, 2021 |
| Switzerland/ZH-ETHZ-500088/2020 | Nov 09, 2020 | Zurich | B.1.1.7 | Viollier AG | D-BSSE, ETH Zurich | Mar 2, 2021 |
| Switzerland/ZH-ETHZ-500086/2020 | Nov 09, 2020 | Zurich | B.1.1.7 | Viollier AG | D-BSSE, ETH Zurich | Mar 2, 2021 |
| Switzerland/ZH-UZH-IMV130/2020 | Dec 18, 2020 | Zurich | B.1.1.7 | University Hospital Zurich | Institute of Medical Virology, University of Zurich | Dec 30, 2020 |
